# Variation among states in reducing coronavirus spread: impact of stay-at-home orders

**DOI:** 10.1101/2020.04.27.20081752

**Authors:** Richard Condit

## Abstract

The corona virus, COVID-19, has been spreading rapidly across the USA since early March, but at a decreasing rate, where the rate r is defined as the exponential increase. I modeled the way the rate of increase *y* = log(exp(r)-1) has declined through time in each of the 51 states with the goal of determining whether state-at-home orders correlate with reductions in the rate of spread of the virus. A piecewise linear regression was used, with a single break point. This model can identify whether there was a change in the rate of decline, when the change happened, and which states have shown the greatest improvement in reducing the spread of COVID-19. The piecewise model identified a significant breakpoint on 24 Mar for all states combined, and all states had nearly the same breakpoint. Prior to 24 Mar, the average change in *y* was -0.013 per day, meaning a reduction in the rate of spread from 23.5 pct. per day to 19.5 pct. per day; after 24 Mar, the average change in *y* was -0.070 per day, a reduction from 19.5 pct. per day to 7.5 pct. per day. Prior to 24 Mar there was no significant variation among states in the decline in y, but after 24 Mar there was substantial variation, and the date on which states issued stay at home orders correlated with that variation. Montana, Idaho, and Vermont showed the greatest improvement, while Nebraska, South Dakota, and Iowa the least. The improvement as measured by the reduction after 24 Mar did not correlate with case density in a state, nor state population.

## 1 Introduction

The number of COVID-19 infections has increased steadily since early March in every state in the USA. The density of infections (cases per capita) varies substantially among states, but the more pertinent interest is the rate at which the number of infections increases. Various public health measures have been taken to slow that increase, and my goal here is to assess one such measure – the stay-at-home order – has impacted the rate of spread. Such assessments have been done to test for the effect of measures taken to combat the 1918 influenza epidemic (Bootsma and Ferguson 2007). The assessment is based on a rigorous comparison of the rate of COVID-19 spread in all states in the USA. Though many factors might affect that rate, a longitudinal comparison within each state should be a way to judge how effective the measures were. States with sooner measures should show steeper declines in the rate of spread through time.

## 2 Materials and Methods

### 2.1 Data assembly

Daily counts of the cumulative total of COVID-19 cases per state was collected from weather.com (weather.com 2020). There is a stable url for each state that I could *curl* (the unix function to capture web text) with an automated script. Each day’s web presentation was complete, including daily counts back to mid-February. A url for all 51 states (with DC) had to be copied and saved, but once stored, capturing all states’ information was a fully automated process. The text came as a long html script with javascript data arrays giving numbers buried within. I wrote C++ program to extract those arrays and move them into tables in the R programming language. Many individual records were checked to confirm the data were captured correctly. Counts were cross-checked against data from a New York Times Github site (NY Times 2020) and were essentially (but not exactly) identical. Analyses were done on case records through 21 Apr 2020, downloaded on 23 Apr 2020. More recent data are shown in graphs, but were not used in the model.

### 2.2 New cases and deaths

The data come as the cumulative number of cases of COVID-19 and cumulative number of deaths attributed to the virus. I calculated the number of new cases per day as the difference between total cases on successive days. In a few cases where no new cases were reported on one day, I used the new cases on the next day divided by the number of intervening days and omitted the day with no reports from analyses. This avoided having any zeroes in the new case counts. As an estimate of current cases, I used the cumulative count each day minus the number of deaths. Ideally, I would have used the number of active cases, substracting also all recoveries, but that information was not available.

### 2.3 Rate of increase

Let the cumulative number of cases on day *t* be *N_t_*, so the rate constant of population growth r is defined from

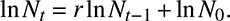

If *r* is constant through time, growth is exponential but I do not make this assumption. The number of new cases on day *t* is *C_t_* = *N_t_* _−_ *N_t_*_−1_ so

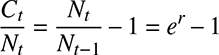

and

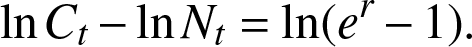

Define *y =* ln(*e^r^* − 1) as the response variable in a model of rate of spread of COVID-19. It has a roughly Gaussian distribution, and it has been changing linearly through time over the past 6 weeks in the US. Note that if *r* ~ 0.2 or less, *e^r^ ~* 1 + *r* so *y* ~ *r*, and *r* is the fractional daily increase. When *r* >> 0.2, *y* increases monotonically with r but is a better choice due to its symmetrical distribution.

### 2.4 Modeling the changing rate of increase

Since the core question is whether there was a shift in the rate of increase as a result of public health measures, I chose a linear piecewise regression (McGee and Carleton 1970) model of y, the rate of increase of COVID-19, versus time. Since the rate *y* describes the change in the total number of cases through time, the model describes an acceleration (or deceleration): *y* is the first derivative, so the change in *y* through time is the second derivative of the number of cases. The following analysis is all about that second derivative, or how the rate of increase changes. If growth were exponential, the rate would not change, ie the second derivative of *N_t_* would be zero. As everyone watching knows, the rate of spread of COVID-19 has been declining, and the model I create here fits that decline as a linear response to time.

The piecewise component of the regression adds the feature that the decline in the rate of spread changes. Consider a piecewise regression with two phases: there is a single breakpoint where the slope of *y* versus *t* shifts, and thus two different slopes, one on either side of the break point. This approach specifically answers the question whether there was a shift in the way the rate changed and whether individual states differed in the shift. I fit the model allowing each state to have a distinct response, so the results can identify states where there was a substantial improvement, that is, a shift toward much slower spread, and other states where the rate steadily declined without any improvement. Alternatively, the model can report no difference states. There is no a priori assumption about when the shift happened – the model will choose the breakpoint based on the data, and the model will report a rigorous test about whether or not there is a break, ie whether the slope changes.

I used a multi-level hierarchical model, also known as a mixed-effects model, in which the 51 states were random effects. This produces an estimate for how the rate of COVID-19 has changed through time in every state, but has the benefit of simultaneously using all the states together. This adds power to the model (Gelman and Hill 2007), important given that there are only 50 days in the analysis and those days must be divided into two phases. Parameters were fitted using a Bayesian approach based on a Gibbs sampler, as detailed in Condit et al. (2007, 2013, 2014). The Bayesian method produces 95 pct. credible intervals based on every parameter estimate examined by the sampler, and I inferred statistical difference between estimates when 95 pct. credible intervals did not overlap. I also tried a three phase piecewise linear model and simple linear regression (ie no break point). The three models were compared using the deviance information criterion (DIC); a lower DIC means a better model fit (Plummer 2008).

### 2.5 Testing the impact of health measures

A Wikipedia article includes measures and dates of implementation in every state (Wikipedia 2020). The measure most clearly offering a quantitative measure across states is the date on which stay-at-home orders were issued. Four states have not issued such orders (as of 2 May), so they were assigned a date of 10 Apr, three days after the latest order (South Carolina, 7 Apr). Three states had various orders locally, and they were excluded. The rate of improvement in each state after 24 Mar, as estimated from piecewise regression, was correlated against the date of the stay-at-home order.

## 3 Results

### Increase in total cases

The number of COVID-19 cases increased steadily but at a consistently declining rate (Fig.1). That is, growth was less than exponential.

**Figure 1.**
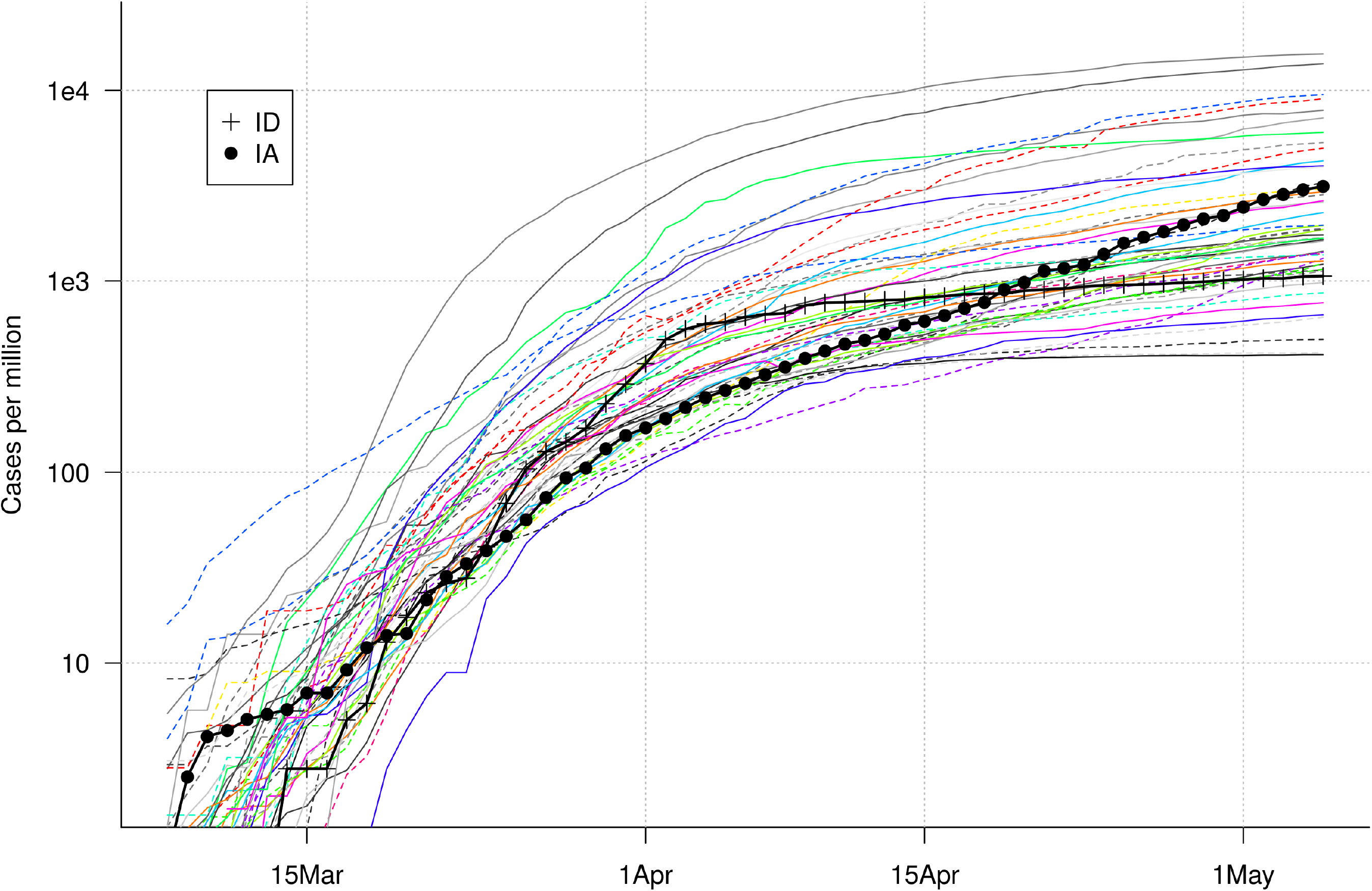
Cumulative number of COVID-19 cases per day in all 51 states on a log-scale. Each state is colored differently. With the vertical axis logged, a constant rate of increase would result in a straight line, so it is clear that the rate has steadily declined in all states. Montana and Nebraska are highlight to compare with Fig. 2. Click figure to enlarge.

### Decrease in daily rate of change

The rate of increase has declined in a roughly linear fashion since early March (Fig. 2). Piecewise regression identified a break on 24 March with strong statistical support. The mean slope of all 51 states (fixed effect of the model) prior to 24 March was −.013, steepening to −.070 after 24 March. Those slopes are equivalent to reducing the rate of spread from 23.3 pct. per day to 19.5 pct. per day for the two weeks prior to 24 Mar, then from 19.5 pct. to 7.5 pct. per day in the two weeks after.

**Figure 2.**
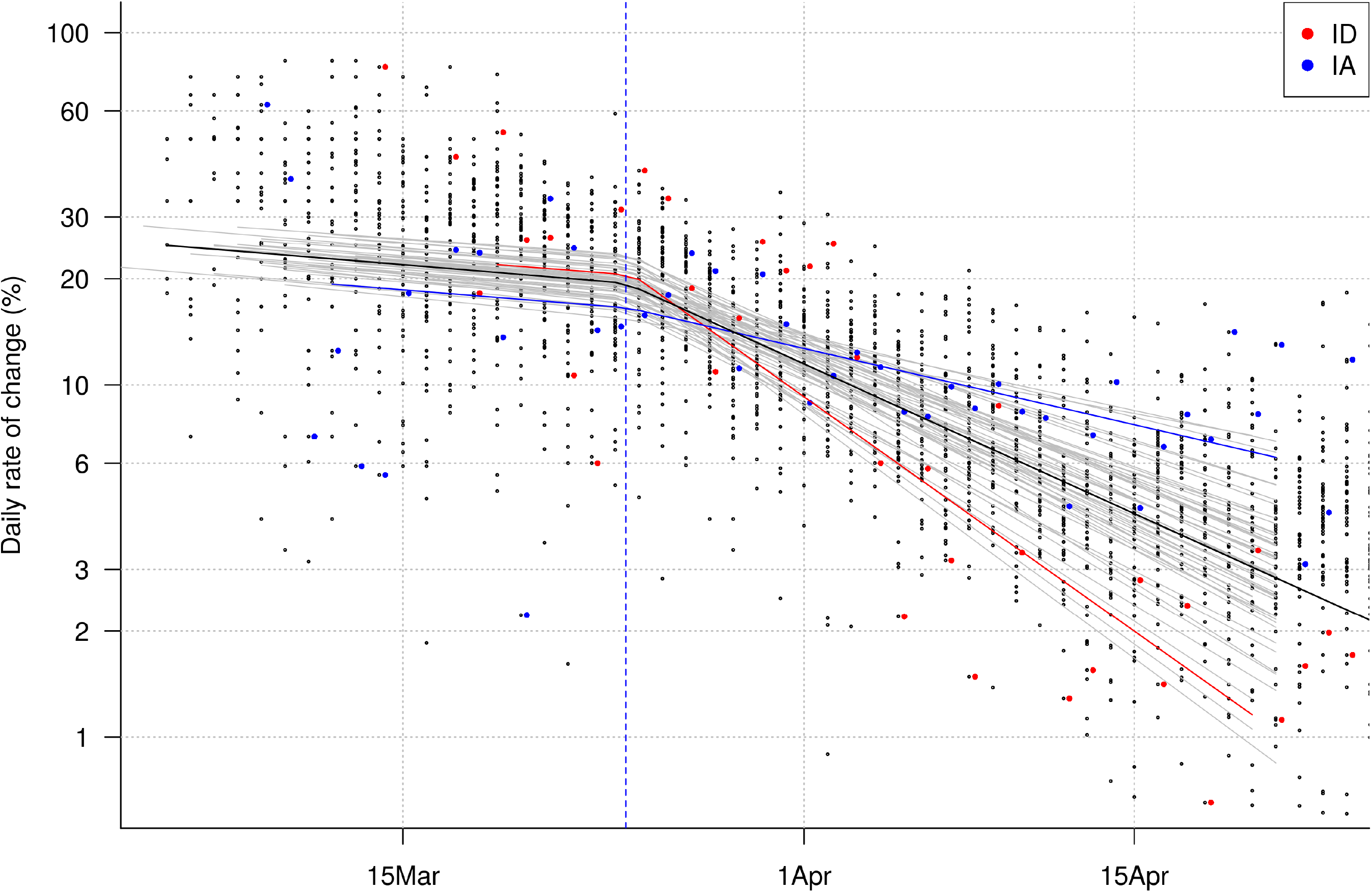
The daily rate of increase of COVID-19 since early March, with *y* = ln(*e^r^* − 1) on the vertical axis (see Methods). Each point is a single daily change in one state. The labels on the vertical axis convert *y* to a percent per day (see Methods). That the rate is always positive means the virus is spreading. The important observation is that the rate at which it is spreading has been declining steadily, with a shift in the decline on 24 March (the vertical blue line). The heavy black lines show the model’s estimate of the mean daily rate (all 51 states). Each fine gray line is the models’ estimate of the rate in individual states. Montana and Nebraska are highlighted as extremes in reducing the rate (Montana) or not (Nebraksa). Compare with Figure 1. Click figure to enlarge.

There was no variation across states in the day on which the slope changed: in all 51 states it was either 24 Mar and 25 Mar, and credible for all 51 states overlapped. Likewise, the slope prior to the break did not vary significantly among states; all 51 credible intervals overlapped, and the slope was always between −0.16 and −.010.

There was, however, statistically significant variaton among states in the slope after 24 Mar. Individual states had slopes varying from −0.114 to −0.029. Idaho illustrates strong improvement, at a rate of −0.109 (Montana was slightly better at −0.114; in Idaho, was from 20.6 pct. per day on 24 Mar to 4.8 pct. per day two weeks later. Iowa illustrates poor improvement, with a rate of −.0356, from 16.6 pct. per day on 24 Mar to 10.3 pct. per day two weeks later. Thus, the virus was spreading faster in Idaho than in Iowa on 24 Mar, but the trend had reversed by 8 Apr. Those two states are highlighted in Fig. 1 and Fig. 2 to illustrate extremes. The slopes in two phases in all 51 states are given in Table 1 and are available for download (see Supplementary Data).

**Table 1.** Improvement in rate of spread of COVID-19 as measured by the slope of the rate of increase through time, divided in two time periods. The break between time periods was estimated as 24 Mar, and slopes are given before and after that break (with 95% credible intervals in parentheses. The slopes and the break were estimate using piecewise regression.

| State | Slope before 24 Mar (95% CI) | Slope after 24 Mar (95% CI) |
| --- | --- | --- |
| AK | -0.01275 (-0.02473,6e-04) | -0.09231 (-0.10922,-0.07729) |
| AL | -0.01249 (-0.02500,0.00079) | -0.06605 (-0.08104,-0.05122) |
| AR | -0.01462 (-0.02849,-0.00305) | -0.07203 (-0.08543,-0.05697) |
| AZ | -0.01077 (-0.02079,0.00405) | -0.07027 (-0.08727,-0.05481) |
| CA | -0.01328 (-0.02349,-0.00233) | -0.07866 (-0.09438,-0.06444) |
| CO | -0.01624 (-0.03189,-0.00491) | -0.06321 (-0.07855,-0.04863) |
| CT | -0.01111 (-0.02093,0.00369) | -0.06853 (-0.08319,-0.05341) |
| DC | -0.01329 (-0.02570,-0.00082) | -0.05479 (-0.07067,-0.03995) |
| DE | -0.01245 (-0.02326,0.00095) | -0.04825 (-0.06436,-0.03195) |
| FL | -0.01269 (-0.02378,0.00078) | -0.07595 (-0.09094,-0.06061) |
| GA | -0.01332 (-0.02369,-0.00207) | -0.06616 (-0.08074,-0.05124) |
| HI | -0.01353 (-0.02735,-0.00091) | -0.10188 (-0.11811,-0.08536) |
| IA | -0.01234 (-0.02358,-0.00035) | -0.03552 (-0.05173,-0.01875) |
| ID | -0.01204 (-0.02345,0.00296) | -0.10943 (-0.12730,-0.09170) |
| IL | -0.01046 (-0.02069,0.00368) | -0.06095 (-0.07637,-0.04582) |
| IN | -0.01113 (-0.02104,0.00266) | -0.06635 (-0.08342,-0.05185) |
| KS | -0.01183 (-0.02158,0.00092) | -0.07172 (-0.08996,-0.05703) |
| KY | -0.01305 (-0.02507,-0.00043) | -0.05857 (-0.07338,-0.04338) |
| LA | -0.01204 (-0.02273,0.00139) | -0.09828 (-0.11332,-0.08351) |
| MA | -0.01131 (-0.02142,0.00325) | -0.05582 (-0.07074,-0.04100) |
| MD | -0.01122 (-0.02094,0.00321) | -0.05546 (-0.07119,-0.04082) |
| ME | -0.01429 (-0.02720,-0.00145) | -0.08316 (-0.09896,-0.06942) |
| MI | -0.01057 (-0.02119,0.00329) | -0.08639 (-0.10140,-0.07114) |
| MN | -0.01533 (-0.03173,-0.00342) | -0.06121 (-0.07587,-0.04659) |
| MO | -0.01107 (-0.02131,0.00209) | -0.07833 (-0.09483,-0.06254) |
| MS | -0.01211 (-0.02274,0.00176) | -0.06151 (-0.07517,-0.04695) |
| MT | -0.01245 (-0.02352,0.00101) | -0.11394 (-0.13159,-0.09812) |
| NC | -0.01251 (-0.02305,7e-04) | -0.06311 (-0.07676,-0.04841) |
| ND | -0.01532 (-0.02890,-0.00296) | -0.04975 (-0.06434,-0.03443) |
| NE | -0.01545 (-0.02947,-0.00442) | -0.02863 (-0.04471,-0.01262) |
| NH | -0.01325 (-0.02472,-0.00142) | -0.06082 (-0.07610,-0.04558) |
| NJ | -0.01094 (-0.02127,0.00671) | -0.07407 (-0.08969,-0.05902) |
| NM | -0.01300 (-0.02371,-0.00017) | -0.05499 (-0.07150,-0.03919) |
| NV | -0.01360 (-0.02569,-0.00165) | -0.07203 (-0.08731,-0.05738) |
| NY | -0.01314 (-0.02452,0.00088) | -0.08478 (-0.09868,-0.06985) |
| OH | -0.01261 (-0.02367,0.00034) | -0.05446 (-0.06857,-0.04008) |
| OK | -0.01280 (-0.02361,-0.00127) | -0.07405 (-0.08933,-0.05955) |
| OR | -0.01314 (-0.02362,-0.00241) | -0.07298 (-0.08774,-0.05891) |
| PA | -0.01077 (-0.02072,0.00426) | -0.06699 (-0.08317,-0.05257) |
| RI | -0.01188 (-0.02150,0.00172) | -0.03836 (-0.05482,-0.02171) |
| SC | -0.01220 (-0.02225,0.00115) | -0.06945 (-0.08577,-0.05087) |

Table 1 (cont.)
| State | Slope before 24 Mar (95% CI) | Slope after 24 Mar (95% CI) |
| --- | --- | --- |
| SD | -0.01263 (-0.02362,-0.00053) | -0.03339 (-0.04699,-0.01857) |
| TN | -0.01225 (-0.02238,0.00199) | -0.08031 (-0.09556,-0.06485) |
| TX | -0.01245 (-0.02311,0.00026) | -0.06245 (-0.07785,-0.04836) |
| UT | -0.01339 (-0.02550,-0.00150) | -0.07099 (-0.08510,-0.05663) |
| VA | -0.01200 (-0.02143,0.00044) | -0.04649 (-0.06252,-0.03172) |
| VT | -0.01275 (-0.02499,0.00064) | -0.11129 (-0.12895,-0.09546) |
| WA | -0.01421 (-0.02722,-0.00215) | -0.07678 (-0.09373,-0.06010) |
| WI | -0.01394 (-0.02695,-0.00164) | -0.07697 (-0.09213,-0.06209) |
| WV | -0.01064 (-0.02078,0.00527) | -0.08651 (-0.10435,-0.07054) |
| WY | -0.01464 (-0.03273,-0.00386) | -0.09323 (-0.10863,-0.07715) |

There was no correlation between the slope prior to 24 Mar and the slope after that day (Supplemental Fig. S1). This is expected given the lack of variation prior to 24 Mar.

### Improvement in phase 2 and stay-at-home orders

There was a significant correlation between the date each state declared a stay-at-home order and the rate of improvement since 24 Mar (Fig. 3). Included in the regression are four states for which there was no stay-at-home order (as of 2 May). The average state which had issued the order on 19 Mar (the earliest) improved, according to the regression, from an increase of 19.5 pct. per day on 24 Mar to 6.5 pct. per day two weeks later. In comparison, in the average state that delayed until 7 Apr (the latest), the estimated improvement was 19.5 pct. per day to 8.6 pct. per day over those two weeks. The impact of the earlier stay-at-home was an improvement of ~ 2 pct. per day over two weeks, relative to waiting 19 days to issue the order.

**Figure 3.**
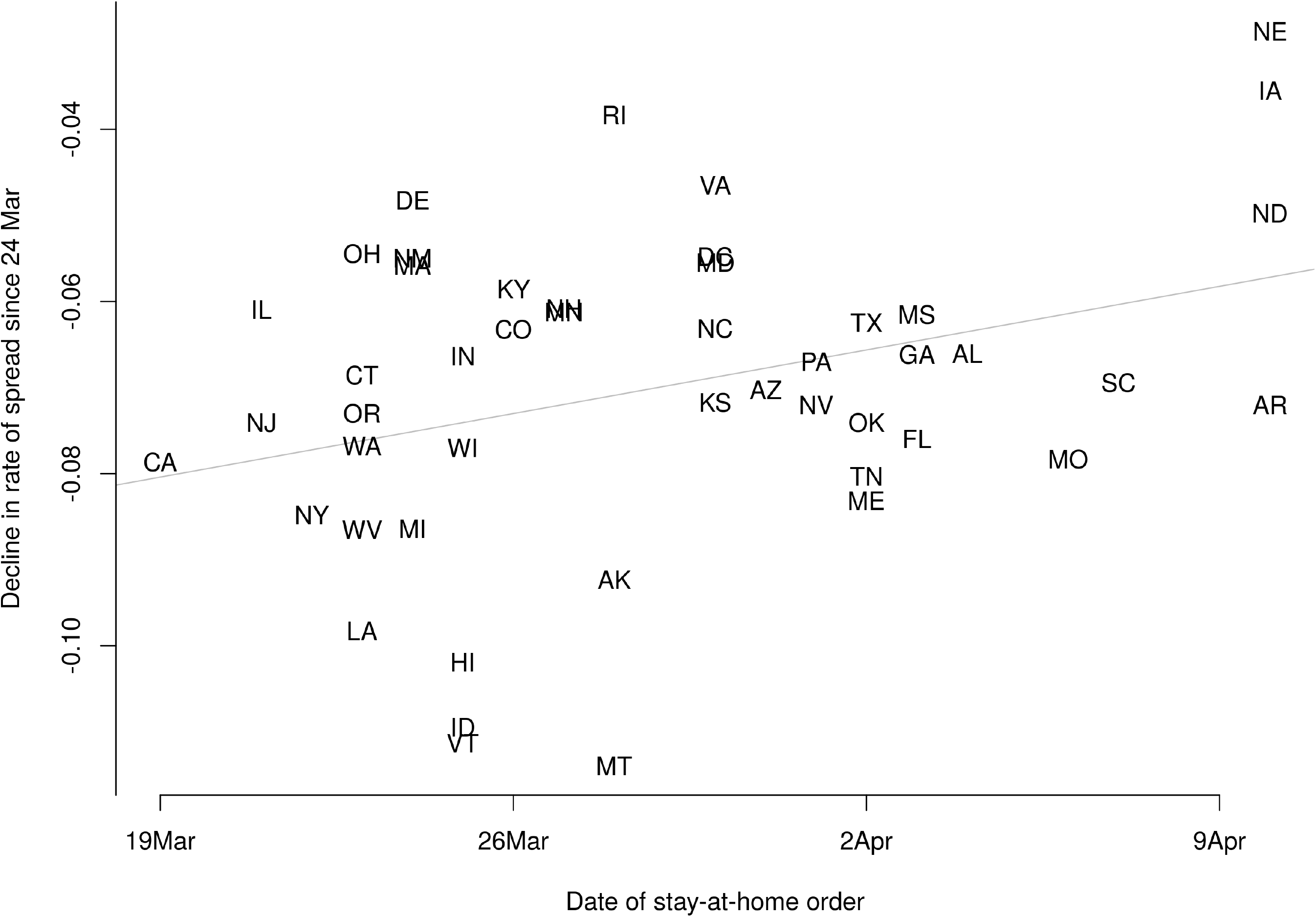
The corona virus, COVID-19, has been spreading rapidly across the USA since early March, but at a decreasing rate, where the rate *r* is defined as the exponential increase. I modeled the way the rate of increase *y* = ln(*e^r^* − 1) has declined through time in each of the 51 states with the goal of determining whether state-at-home orders correlate with reductions in the rate of spread of the virus. A piecewise linear regression was used, with a single break point. This model can identify whether there was a change in the rate of decline, when the change happened, and which states have shown the greatest improvement in reducing the spread of COVID-19. The piecewise model identified a significant breakpoint on 24 Mar for all states combined, and all states had nearly the same breakpoint. Prior to 24 Mar, the average change in *y* was −0.013 *d*^−1^, meaning a reduction in the rate of spread from 23.5 pct. *d*^−1^ to 19.5 pct. *d*^−1^; after 24 Mar, the average change in *y* was −0.070 *d*^−1^, a reduction from 19.5 pct. *d*^−1^ to 7.5 pct. *d*^−1^. Prior to 24 Mar there was no significant variation among states in the decline in *y*, but after 24 Mar there was substantial variation, and the date on which states issued stay at home orders correlated with that variation. Montana, Idaho, and Vermont showed the greatest improvement, while Nebraska, South Dakota, and Iowa the least. The improvement as measured by the reduction after 24 Mar did not correlate with case density in a state, nor state population.

**Figure 4.**
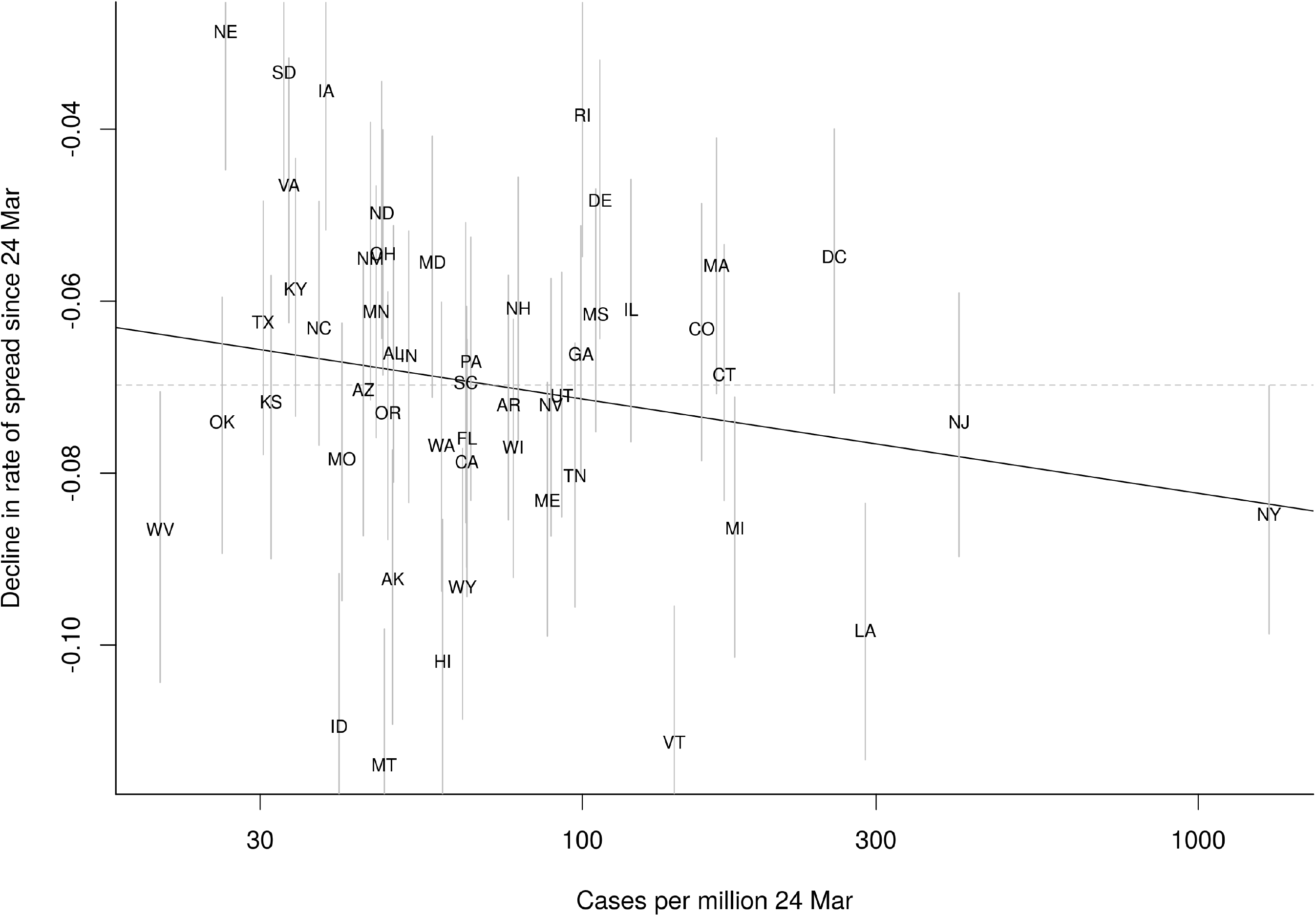
Correlation between the improvement in the rate of spread since 24 Mar (vertical axis) and the case density (per million population) in each state. A negative on the y-axis means declining (ie improving) rate of spread, so the more negative values, toward the bottom of the graph, had the greatest improvement. The regression is not significant (*p* = 0.17, *r*^2^ = 0.04). Though New York, with the highest case density, had better than average improvement, states near the middle in case density had a wide range of improvement, from Idaho, Montana, Hawaii, and Vermont with great improvement to Virginia, Iowa, Rhode Island, and Delaware with little improvement.

### Improvement in phase 2 and case density

There was no correlation between the improvement in the rate of spread, as measured by the slope after 24 Mar in each state, and the case density (cases per million) on 24 Mar (Fig. 3). The slope was more negative (better improvement) in states with a higher density of cases, but the regression was not significant (Fig. 3).

### Improvement in phase 2 and population density

There was no correlation between the improvement in the rate of spread, as measured by the slope after 24 Mar in each state, and the population size of a state. The slope was slightly positive but non-significant (*p =* 0.63, *r*^2^ ~ .01).

### Alternative models

Three-phase piecewise regression identified one break matching the sharp shift of the two-phase model, plus a later break that was accompanied by no change in slope. Based on the deviance information criterion (DIC), the two phase model (DIC=2392.9) was superior to the three-phase (DIC=2636.0) or a simple regression, with constant slope throughout (DIC=2766.0).

## 4 Discussion

The main conclusion is that the rate at which the corona virus has been spreading across the US changed in different ways in different states. The observation that the rate declined through time – meaning growth is less than exponential – could be attributed to many different factors. But the fact that states differed in the degree of improvement, that is how much the rate declined, must be attributed to differences among states. How much improvement states showed could not be attributed to the density of infections around 24 March, nor to the population size of the state.

The rate of improvement was predicted, however, by the date on which dates issued stay-at-home orders. Three of the states with the poorest improvement, Nebraska, Iowa, and North Dakota, do not have stay-at-home orders (as of 2 May), and several states with the best improvement had early orders (Louisiana, Hawaii, Idaho, Vermont). Issuing the order three weeks earlier led to a reduction in the rate of spread of COVID-19 by about 2 pct.: the average state issuing an early order reduced the rate of spread to 6 pct. per day, while in those with a late order, it improved to 8 pct. per day. But the correlation was not strong, and there are many exceptions, particularly cases with early orders but poor improvement, especially Delaware and Ohio. An analysis of of health measures taken during the 1918 flu pandemic also found moderate and variable impacts (Bootsma and Ferguson 2007).

I would suggest that rates of spread of the corona virus should be modeled using the number of active cases, since those are where new infections arise. But the information available now includes only the cumulative number of cases. Successive cumulative counts yield an accurate estimate of new cases per day, but the denominator of the rate is cumulative cases. I subtracted deaths from that to get closer to active cases, and I am working on estimating the number of recoveries using data on time to recovery (Verity et al. 2020).

Other policy measures have been taken to inhibit COVID-19 spread, and some of these undoubtedly correlate with the stay-at-home order. Which measures are most important remains untested. Many other factors must be impacting how quickly the rate of spread has slowed, and further work should examine other predictors of state-level variation. Testing other predictors with county-level variation might be more effective, and I plan to extend the piecewise regression model presented here to counties.

## Data Availability

Data and software are available for download.

http://conditdatacenter.org/covid19

## 6 Acknowledgments

Alexander Schapiro helped collect web data.

## 7 Supplementary Data

Data and software can be downloaded from the author’s web page, http://conditdatacenter.org/covid19.

**Figure S1.**
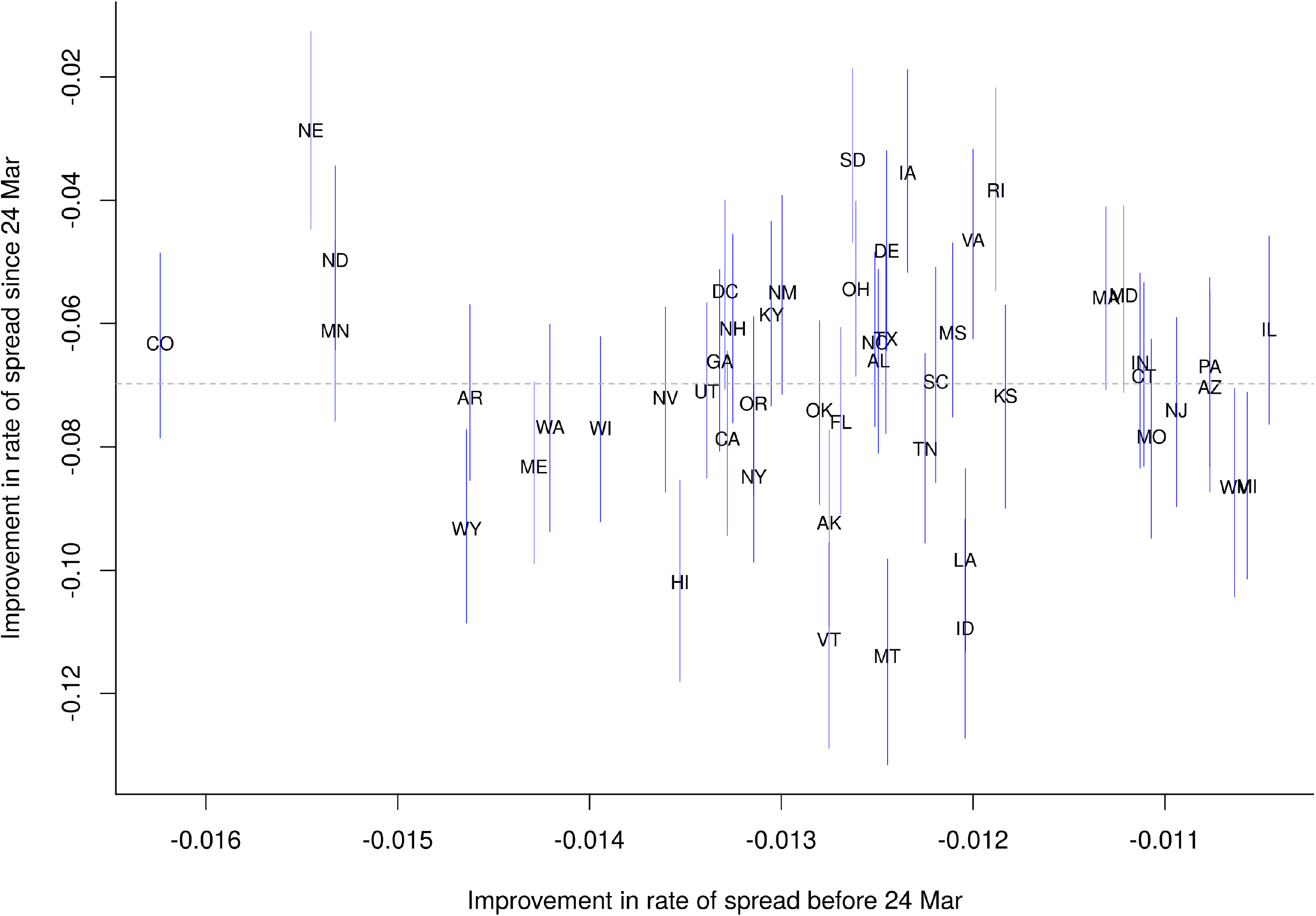
Improvement in rate at which COVID-19 spread in each state prior to 24 Mar (horizontal axis) versus after 24 Mar (vertical axis). Thin vertical bars show 95 pct. credible intervals on the second slope estimates; pairs of states whose vertical bars do not overlap are inferred to be statistically distinct. The horizontal axis has a much narrower range, since states barely differed; horizontal credible bars are omitted because every one would extend outside the range of the figure. There was not a significant correlation between the two slopes.

**Figure S2.**
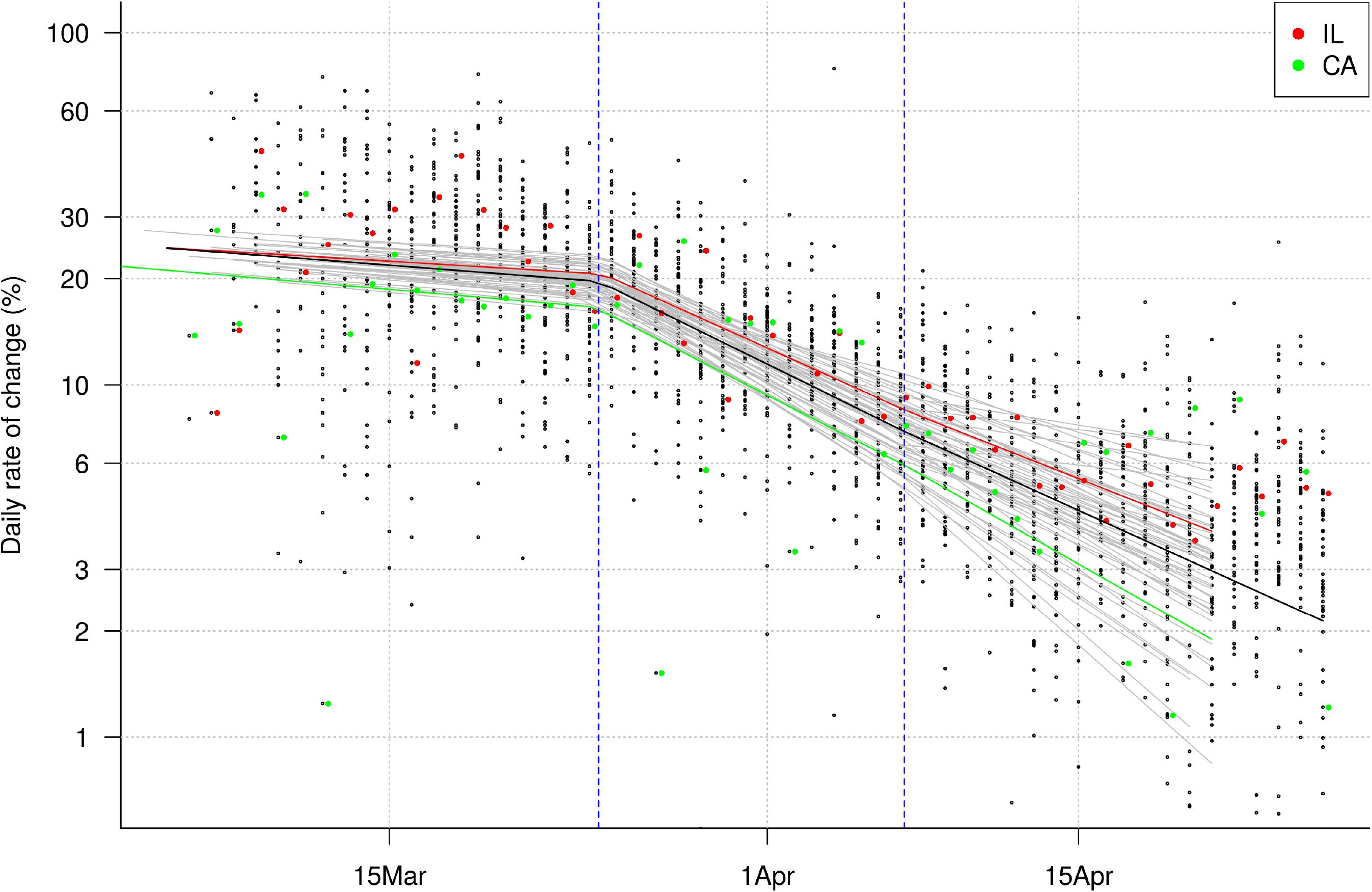
Fit of three-phase piecewise regression to the improvement in rate of spread of COVID-19. See Fig. 2 in main text. The two vertical dashed lines show the two breaks. Slopes of second and third phases did not differ significantly, and the model fit (from DIC) was inferior to the two-phase model. Two different states are highlighted, Illinois and California.

